# Memory and Concentration Skills In A Sample of First Grade Medical Students at University of Baghdad/College of Medicine

**DOI:** 10.1101/2022.04.16.22273927

**Authors:** Zaher Mohammed Fadhil, Mohammed Saad Khashea, Ali Fadhil Abdulqader, Ali M. Jawad Almothaffar

## Abstract

**Objectives:** The purpose of this study is to assess the level of memory skills and concentration skills among first year medical students in College of Medicine/University of Baghdad depending on global scale (Study Skills Inventory SSI).

**Subjects and Method:** A cross-sectional study to assess memory and concentration skills among first year medical students in College of Medicine/ University of Baghdad, the study was conducted using an online survey in September 2020. A sample of 103 students participated in the study by filling of an online questionnaire which was modified from the Study Skills Inventory (SSI).

Regarding memory skills a score less than 30 was considered not adequate, while regarding concentration skills a score less than 35 was considered not adequate.

**Results:** Percentage of males was 68% and the percentage of females was 32%. About studying hours we found that 59.2% students study less than 3 hours, 25.2% students study between 3-6 hours and 15.5% students study more than 6 hours. The mean score of the students for concentration skills was 36.45 and was 32.40 for memory skills. Regarding concentration skills 35% students had non adequate score and 65% students had adequate score, regarding memory skills 28.2% students had non adequate score and 71.8% students had adequate score. There is a statistically significant association between concentration skills and studying hours and there is statistically significant association between memory skills and studying hours. There was a statistically significant moderate positive correlation between concentration skill score and the memory skill score of the students total score (r = 0.511, p < 0.01).

**Conclusion:** About 75% of 1^st^ year medical students have adequate concentration and memory skills. The students who study for 3-6 hours daily have the least mean score of both skills with 42.3% of them have adequate concentration skills score and 57.7% had adequate memory skills score. Further studies with larger sample size are needed to correlate the concentration and memory skills with student end year average total score.

## Introduction

The transition from relatively stable and didactic teaching at high school to the strikingly different reality of medical school is challenging for medical students worldwide. The significant amount of educational content leave many students in frustration, failure, and psychological morbidities like stress, depression, and anxiety. It is often stated that study skills that made students successful in high school turn out quite insufficient and inefficient for success in medical school .^(1,2)^

Academic performance of students is one of the main indicators used to evaluate the quality of education in universities .^(3,4)^ Academic performance is a complex process that is influenced by several factors, such as study habits. ^(4)^ Study habit is a diverse individual behavior in relation to studying, and is a combination of study methods and skills .^(5,6)^ In other words, study habits include behaviors and skills that can increase motivation and convert the study into an effective process with high returns, which ultimately increases the learning. ^(7)^ This skill is also defined as any activity that facilitates the process of learning about a topic, solving the problems or memorizing part or all of the presented materials. ^(5)^

Students realize that the overwhelming burden of information needs to be addressed and requires certain coping strategies. It is also clear that those who face difficulty in coping come under huge academic pressure, resulting in their poor academic performance .^(8)^

Since it is a difficult task to learn and retain a considerable volume of up-to-date and specialized information the students confront in an academic year which takes them a lot of time and requires them to have planning, it is obvious that lack of appropriate study skills and habits not only leads to the students’ waste of time, energy and their tendency toward bad study habits, but also results in their backwardness which in turn causes confusion and anxiety. ^(9)^

Concentration is the ability to focus and give undivided attention on a single task by ignoring all other distractions. As per Hartley and Davis, average attention span of adult is 10-15 minutes. Observation of environment takes place through immediate concentration. Reading, learning and thinking take place through prolonged concentration.^(10)^ Concentration will help the students to complete the task in shorter period and helps to reduce error.^(11)^ Cell phones are one of the attractive devices that can affect students’ attention and concentration, as students can be easily distracted by text messages and feel the urge to reply instantly .^(12)^

Distraction is one of the most common problems of lack of concentration; it is difficult to keep the concentration of students at its maximum potential during the entire time of the class.^(13)^

Memory is an essential part of the studying skills which students will need in their transition from their schools where they have a few and limited information to the university where there are lots of sources and books which can be almost unlimited. Deficits in daytime performance due to sleep loss are experienced universally and associated with a significant social, financial, and human cost. Microsleeps, sleep attacks, and lapses in cognition increase with sleep loss as a function of state instability. Sleep deprivation studies repeatedly show a variable (negative) impact on mood, cognitive performance, and motor function due to an increasing sleep propensity and destabilization of the wake state. Specific neurocognitive domains including executive attention, working memory, and divergent higher cognitive functions are particularly vulnerable to sleep loss.^(14)^

The purpose of this study is to assess the level of memory skills and concentration skills among first year medical students in College of Medicine/University of Baghdad depending on global scale (Study Skills Inventory SSI).

## Patients and Methods

A cross sectional study conducted on first year medical students currently enrolled in College of Medicine/ University of Baghdad in the Academic Year 2020-2021 who accept to participate in completing the survey.

### Data collection

Participants were asked to fill an online questionnaire that was published using Google forms. The questionnaire, which was modified from the Study Skills Inventory (SSI), There were 26 questions which were divided into:

1. Basic statistical data: A seven-item questionnaire was used to gather information about demographic data (age, gender and residency), studying hours and sleeping hours.
2. Memory and concentration skills: A 19-item questionnaire was used to assess memory skills (9 questions) and concentration skills (10 questions). Questions were based on the Study Skills Inventory (SSI) .^(15)^ This inventory was found to be both reliable and valid in previous studies among medical students.^(16, 17)^

The maximum total score of memory skills is 45 and of concentration skills is 50. Each statement has five options: always, usually, sometimes, rarely and never.

This five-point Likert scale is scored from 1 to 5; i.e.5 stands for always (i.e. all the time - around 100% of the time), 4 stands for usually (i.e. most of the time - around 75% of the time), 3 stands for sometimes (i.e. occasionally- around 50% of the time), 2 stands for rarely (i.e. few times - around 25% of the time), 1 stands for never (i.e. almost at no times - around 0% of the time).

The students were divided into two major groups, termed as adequate and not adequate based on their scores in the study skill inventory, University of Central Florida, regarding memory skills a score less than 30 which is termed as not adequate, regarding concentration skills a score less than 35 .^(15)^

### Ethical Consideration

The approval of the College of Medicine/ University of Baghdad was taken to conduct this research. The online questionnaire explained the purpose of the questionnaire and of the research, also there was a field explaining that participation in filling this survey will be considered as a full consent. Participation was fully optional and the identities of the participants are anonymous.

### Statistical Analysis

The Statistical Package for the Social Sciences (SPSS, version 20) was used for the data analysis. Descriptive statistics such as frequency, percentages, mean, and standard deviation were calculated.

Comparisons were made using Chi square test to compare the categorical variables. ANOVA was used for comparisons with variables having more than two categories. Additionally, correlations were computed using Pearson’s correlation coefficient between the memory scores and concentration scores. This test and any other tests will be used; will be performed with a confidence level of 95% with significant level of P < 0.05.

## Results

A total of 103 respondents, all of them are first grade medical students at University of Baghdad/ College of Medicine. Most of them (60.2%) are 19 years old and (24.3%) of them are 18 years old, the rest of them are 17, 20, 21 or 22 years old.

The percentage of male participation is more than that of females, where the number of males is 70 (68.0%) and the number of females is 33 (32.0%).

We divided participants into 2 groups according to their residency: 87 participant live in Baghdad and 16 participant live in other governorates.

We asked the students about how many hours they typically study in one day and they answered: 61 participants (59.2%) study less than 3 hours a day, 26 participants (25.2%) study between 3-6 hours a day and 16 participants (15.5%) study more than 6 hours a day, (Figure 1).

**Figure 1:**
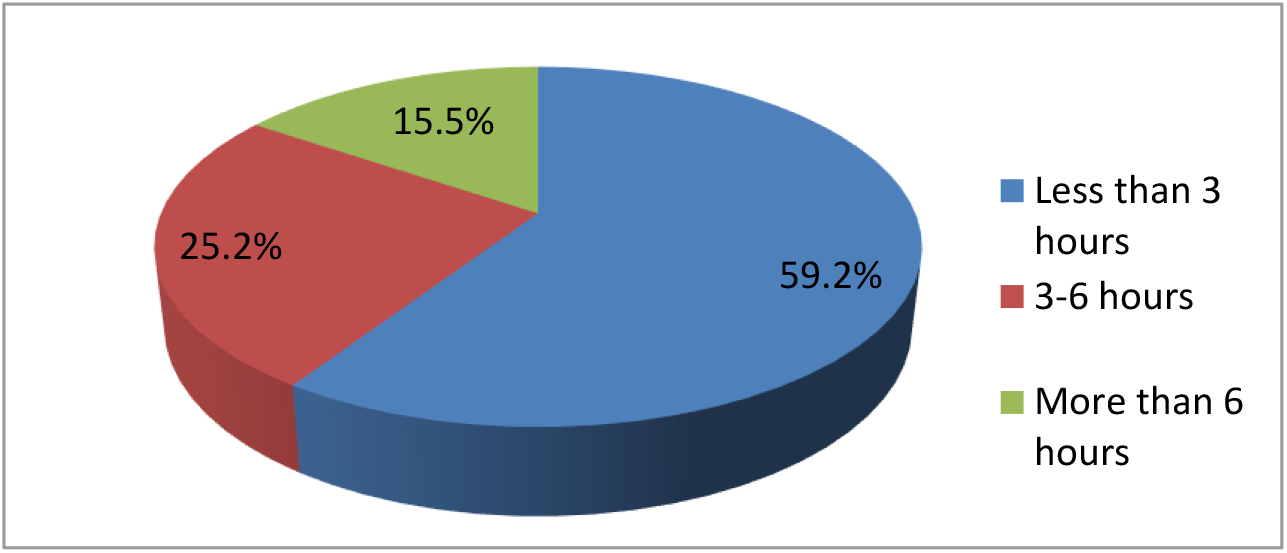
The number of studying hours of the participants.

We asked the students about how many hours they typically sleep in one day and they answered: 62 participants (60.2%) sleep less than 6 hours a day, 12 participants (11.7%) sleep between 6-8 hours a day and 29 participants (28.2%) sleep more than 8 hours a day.

The mean score of the students for concentration skills was 36.45 and was 32.4 for memory skills.

For concentration skills the maximum score of the students was 49 and the minimum score was 24 for memory skills the maximum score of the students was 45 and the minimum score was 14, (Table 1).

**Table 1:** Descriptive data of concentration and memory skills.

|  | CONCENTRATION | MEMORY |
| --- | --- | --- |
| **Mean** | 36.45 | 32.40 |
| **Std. Deviation** | ± 5.376 | ± 5.687 |
| **Minimum** | 24 | 14 |
| **Maximum** | 49 | 45 |

The students were divided into two major groups, termed as adequate and not adequate based on their scores in study skill inventory (SSI), from University of Central Florida, regarding concentration skills a score less than 35 considered as not adequate and equal or more than 35 considered as adequate, 36 participants (35%) had not adequate, 67 participants (65%) had adequate score. (Table 2)

**Table 2:** Concentration and memory Scores of the Study Group.

| Skill | Score | Number | % |
| --- | --- | --- | --- |
| **Concentration** | NOT adequate (<35) | 36 | 35.0 |
|  | Adequate(≥35) | 67 | 65.0 |
|  | **Total** | **103** | **100.0** |
| **Memory** | NOT adequate (<30) | 29 | 28.2 |
|  | Adequate( ≥ 30) | 74 | 71.8 |
|  | **Total** | **103** | **100.0** |

The students were divided into two major groups, termed as adequate and not adequate based on their scores in study skill inventory (SSI), from University of Central Florida, regarding memory skills a score less than 30 considered as not adequate and equal or more than 30 considered as adequate, 29 participants (28.2%) had not adequate., 74 participants (71.8%) had adequate score, (Table 2).

Table 3 summarizes the association of some parameters with the concentration and memory skills.

**Table 3:**
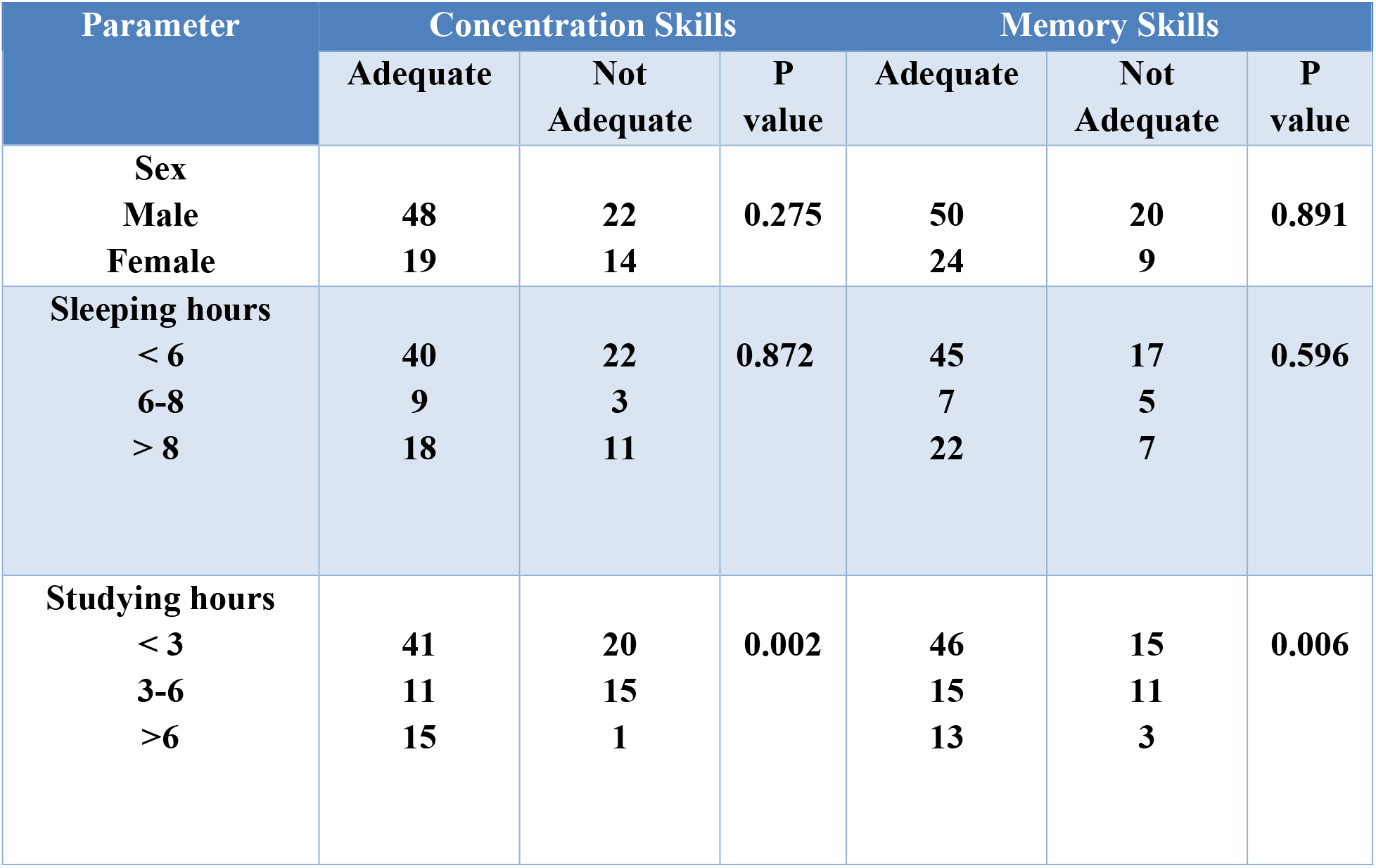
Association of concentration and memory skills with certain factors.

Regarding association of gender and concentration skills,48 (68.6%) of males had adequate score and 22 (31.4%) had not adequate score and about 19 (57.6%) of females had adequate score and 14 (42.4%) had not adequate score. There was no significant association between gender and concentration skills (P value=0.275). Regarding association of gender and memory skills, 50 (71.4%) of males were adequate and 20 (28.6%) were not adequate and about 24 (72.7%) of females were adequate and 9 (27.3%) were not adequate.

There was no significant association between gender and memory skills (P value=0.891).

Regarding association of sleeping hours and concentration skills, 40 (64.5%) of students who sleep less than 6 hours had adequate and 22 (35.5%) had not adequate score. Nine (75%) of students who sleep 6-8 hours had adequate and 3 (25%) had not adequate score. Eighteen (62.1%) of students who sleep more than 8 hours had adequate and 11 (37.9%) had not adequate score. There was no significant association between Sleeping hours and concentration skills.

Regarding association of sleeping hours and memory skills, 45 (72.6%) of students who sleep less than 6 hours had adequate and 17 (27.4%) had not adequate score. Seven (58.3%) of students who sleep 6-8 hours had adequate and 5 (41.7%) had not adequate score. Twenty two (75.9%) of students who sleep more than 8 hours had adequate and 7 (24.1%) had not adequate score.There was no significant association between Sleeping hours and memory skills.

Regarding the association of studying hours and concentration skills, 41 (67.2%) of students who study less than 3 hours had adequate and 20 (32.8%) had not adequate score. Eleven (42.3%) of students who study 3-6 hours were adequate and 15 (57.7%) were not adequate. Fifteen (93.8%) of students who study more than 6 hours were adequate and 1 (6.2%) was not adequate.

There is a statistically significant association between studying hours and concentration skills.

There was a statistically significant moderate positive correlation between concentration skill score and the memory skill score of the students total score (r = 0.511, p < 0.01),Figure 2.

**Figure 2:**
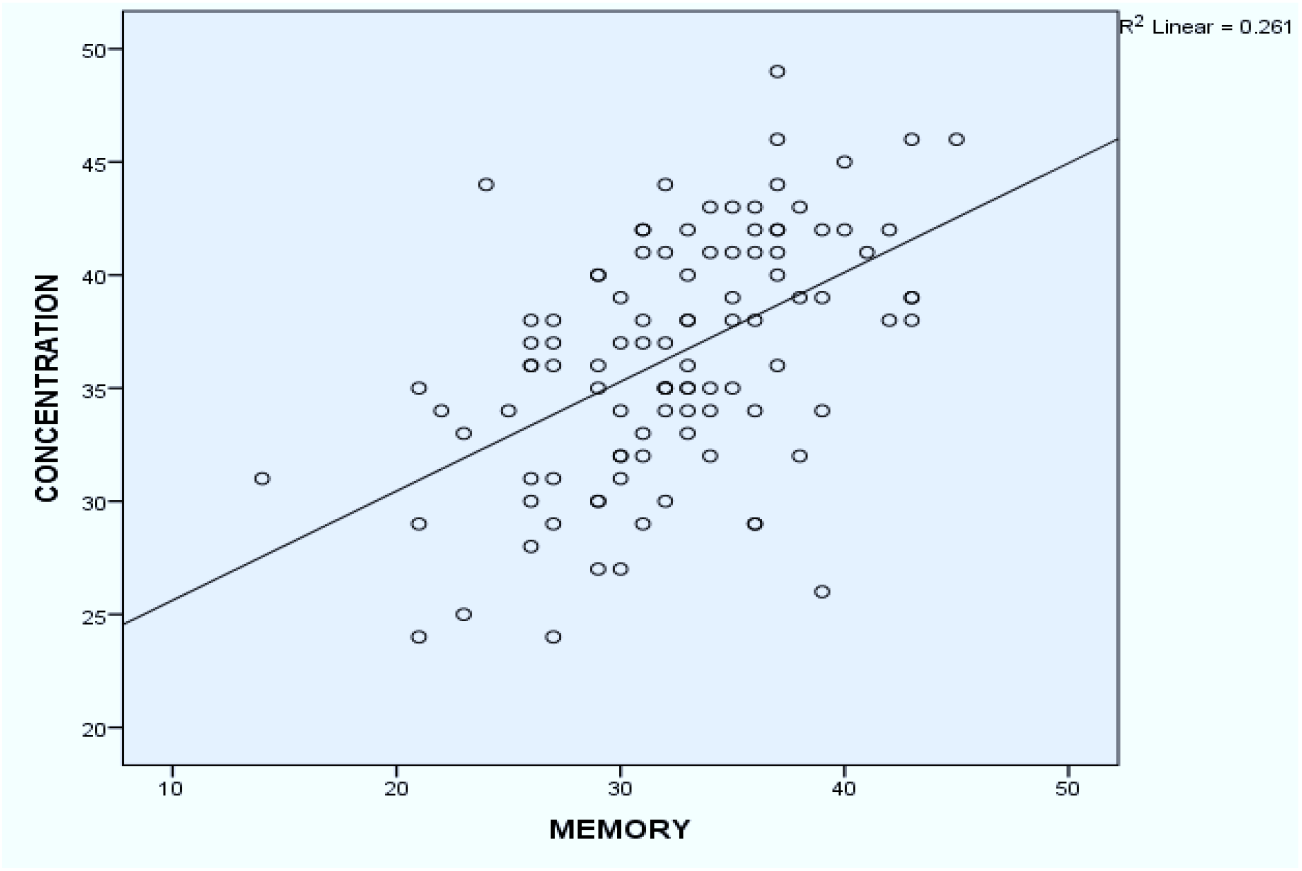
Correlation between concentration skill score and the memory skill score.

## Discussion

In this cross-sectional study which is conducted for 103 first grade students of Baghdad medical college, 70 of the total participants were males and 33 were females.

According to sleeping hours we found that most students (60%) sleep less than 6 hours a day which is less than recommended.^(17)^ Students should change their sleeping pattern and raise their sleeping duration according to Study in Prevalence of Healthy Sleep Duration among Adults — United States, 2014. To promote optimal health and well-being, adults aged 18-60 years are recommended to sleep at least 7 hours each night.^(18)^ Sleeping <7 hours per night is associated with increased risk for obesity, diabetes, high blood pressure, coronary heart disease, stroke, frequent mental distress, and all-cause mortality.^(19, 20)^

This study revealed that 68.6% of total males had adequate, and 57.6% of total females had adequate concentration skills, regarding the memory skills, this study revealed that 71.4% of total males had adequate and about 72.3% of total females had adequate memory skill scores. It seems that both genders have comparable capabilities. The same results had already been obtained by Hossaini et al.^(21)^ In another study by Nourian, the male students had lower scores on comprehension, concentration while studying and time management than females one. The mean score in that study for the concentration skills of males was 2 and for females was 2.25 and difference between the two genders was statistically significant (P value=0.02).^(22)^

Regarding the sleeping hours and concentration skills this study revealed that 64.5% of 62 participants who sleep less than 6 hours had adequate score & 75% of 12 students who sleep 6-8 hours had adequate score and 62.1% of 29 participants who sleeps more than 8 hours had adequate score. This is consistent with other studies.^(23)^

Regarding the sleeping hours and memory skills this study revealed that 72.6% of 62 participants who sleep less than 6 hours had adequate score, 58.3% of 12 participants who sleep 6-8 hours had adequate score and 75.9% of 29 participants who sleep more than 8 hours had adequate score. However, there was no significant association between sleeping hours and memory and concentration skills.

Other studies concluded that shorter acute free-living sleep and sleep deprivation may negatively affect difficult memory tasks, however the relationship between free-living sleep and cognitive task performance in healthy adolescents is less clear than that of laboratory findings, perhaps due to high night-to-night sleep variation and delayed sleep schedule during weekends was associated with poor academic performance. The associations were somewhat reduced after additional adjustment for non-attendance at school, but remained significant in the fully adjusted models. ^(24, 25, 27)^

Regarding studying hours and concentration skills, this study reveals that 67.2% of 61 participants who study less than 3 hours had adequate score, while 42.3% of 26 participants who study 3-6 hours had adequate score and 93.8% of 16 participants who study more than 6 hours had adequate score. There was a statically significant association between studying hours and mean score of concentration skills (p = 0.002). Participants who study 3-6 hours had less concentration skills with mean of scores 33.65 compared with those who study less than 3 hours who had intermediate concentration skills with mean of scores 36.85 and those who study more than 6 hours who had best concentration skills with mean of scores 39.44.

Regarding studying hours and memory skills, this study revealed that 75.4% of 61 participants who study less than 3 hours had adequate score, while 57.7% of 26 participants who study 3-6 hours had adequate score and 81.2% of 16 participants who study more than 6 hours had adequate score. There was a statistically significant association between studying hours and memory skills score (p = 0.006). Participants who study 3-6 hours had less memory skills with mean of scores 30.27 compared with those who study less than 3 hours who had intermediate memory skills with mean of scores 32.38 and those who study more than 6 hours who had best memory skills with mean of scores 35.94.

It seems that students who study 3-6 hours have the least concentration and memory skills; their total number was 26 students (25.2% of total participants). According to their gender: 14 students were males and 12 students were females. As for their sleeping hours: 11 students sleep less than 6 hours a day, 4 students sleep 6-8 hours a day and 11 participants sleep more than 8 hours a day. The details of their sleeping pattern like timing and consistency need to be determined specially for those who have sleeping less than 6 hours and more than 8 hours.

Ukpong and George in their research concluded that academic performance of the long study time behaviour students was significantly different from that of their short study time counterparts. and they recommended that students should set a study time table long enough for effective academic exercises (at least two to three hours daily) for their private study and stick to it.^(27)^

Bin Abdulrahman et al concluded that there is significant correlation between study habits and students’ academic accomplishments. The top ten study habits of highly effective medical students usually have daily study hours ranging between 3 and 4 hours.^(28)^

Regarding correlation between memory and concentration skills the study revealed that there was a statistically significant moderate positive correlation between concentration skill score and memory skill score of the students total score, that means students who got high concentration skills score, got also high memory skills score.

In conclusion, 75% of 1^st^ year medical students have adequate concentration and memory skills. The students who study for 3-6 hours daily have the least mean score of both skills with 42.3% of them have adequate concentration skills score and 57.7% had adequate memory skills score. Further studies with larger sample size are needed to correlate the concentration and memory skills with student end year average total score.

## Limitations

Only 103 individuals were studied, which was a limitation as it is a smaller group of participants. Only students attending one college were studied which limits the diversity of the answers as it was a limited population of students. The data was only being collected online because of the pandemic, which was a limitation.

## Data Availability

All data produced in the present study are available upon reasonable request to the authors

